# EVALUATION OF VITAMIN A SUPPLEMENTATION AND OTHER INTERVENTIONS COVERAGE DURING MATERNAL AND CHILD HEALTH WEEK

**DOI:** 10.1101/2025.06.30.25330575

**Authors:** Mordecai Oweibia, Tarimobowei Egberipou, Gift Cornelius Timighe, Ebiakpo Agbedi, Zuofa Seimo Egberipou, Nkechi Martha Johnson

## Abstract

**Introduction:** Vitamin A Deficiency (VAD) remains a significant public health challenge, particularly in low- and middle-income countries, contributing to childhood morbidity and mortality. In Nigeria, the Maternal, Newborn, and Child Health (MNCH) Week serves as a critical platform for delivering interventions like Vitamin A Supplementation (VAS), deworming, and malnutrition screening. Despite its institutionalization, gaps in coverage, equity, and integration persist. This study evaluated the coverage and effectiveness of VAS and associated interventions during the June 2025 MNCH Week in Bayelsa State, Nigeria, to identify implementation strengths and challenges.

**Methods:** A descriptive cross-sectional design was employed, utilizing secondary data from the official MNCH Week OPS Room Report. The study analyzed coverage levels for VAS, deworming, Iron-Folate supplementation, malnutrition screening (MUAC), HPV vaccination, birth registration, and routine immunization across eight Local Government Areas (LGAs) in Bayelsa State. Data were extracted from validated registers, service summary sheets, and mobile reports, with coverage calculated as the percentage of targeted populations reached. Descriptive statistics were used to summarize findings, and performance trends were visualized using tables and graphs.

**Results:** The study revealed an 83% coverage rate for VAS, exceeding the WHO-recommended threshold of 70%. Deworming coverage lagged at 55%, while Iron-Folate supplementation reached 2,197 pregnant women. Malnutrition screening identified 118 cases of severe acute malnutrition (Red MUAC). HPV vaccination coverage was anomalously high at 330%, whereas birth registration remained critically low at 1%. Routine immunization delivery was inconsistent, particularly in hard-to-reach areas. Disparities in service uptake highlighted logistical and operational challenges, including supply chain gaps and inadequate community mobilization.

**Conclusion:** The MNCH Week achieved high VAS coverage and identified critical malnutrition cases, demonstrating its potential for scalable child health interventions. However, uneven performance in deworming, birth registration, and routine immunization underscores systemic weaknesses. Recommendations include strengthening supply chains, improving data accuracy, enhancing intersectoral collaboration, and prioritizing equity in service delivery. These findings contribute to evidence-based strategies for optimizing campaign effectiveness and advancing child health outcomes in similar settings.

## 1.0 INTRODUCTION

### 1.1 Background of the Study

Vitamin A Deficiency (VAD) remains a pressing global public health issue, particularly affecting low- and middle-income countries where malnutrition, poverty, and limited access to healthcare intersect. Vitamin A is a fat-soluble micronutrient essential for maintaining healthy vision, immune function, epithelial integrity, and overall growth and development. Its deficiency in early childhood increases susceptibility to infections, impairs recovery, and substantially elevates the risk of mortality from common illnesses such as measles and diarrheal disease.

Globally, it is estimated that hundreds of millions of children under the age of five suffer from VAD, with the highest burden in sub-Saharan Africa and South Asia. In response, international health agencies such as the World Health Organization (WHO) and UNICEF recommend high-dose Vitamin A Supplementation (VAS) every 4–6 months for children aged 6 to 59 months in areas with moderate to severe deficiency. VAS has been shown to reduce child mortality by up to 24% in deficient populations, making it one of the most cost-effective and scalable public health interventions available.

In Nigeria, VAD continues to contribute significantly to childhood morbidity, blindness, and mortality, particularly in rural and hard-to-reach areas where access to routine healthcare services is sparse. According to national policy frameworks, including the National Guidelines on Micronutrient Deficiencies Control (Federal Ministry of Health Nigeria, 2020), VAS is recommended semi-annually for all children aged 6–59 months. The implementation is carried out either through routine primary health care facilities or via campaign-based models, such as the biannual **Maternal, Newborn and Child Health (MNCH) Week**.

MNCH Week is a strategic nationwide initiative designed to integrate the delivery of essential child survival interventions. During these weeks, services such as Vitamin A supplementation, deworming (Albendazole), immunization, nutrition screening using MUAC (Mid-Upper Arm Circumference), antenatal micronutrient supplementation, and birth registration are bundled and delivered as a one-stop package, especially targeting underserved communities. The aim is to scale up equitable access, boost coverage figures, and align national progress with Sustainable Development Goals (SDGs), particularly SDG 3 (Good Health and Well-being) and SDG 2 (Zero Hunger).

Despite the institutionalization of MNCH Week as a delivery platform, challenges persist. Reports over the years have documented inconsistencies in service coverage, inequitable delivery between urban and remote LGAs, data quality gaps, weak integration with civil registration systems, and delayed mobilization of resources. There have also been repeated concerns around the sustainability of campaign-based service delivery versus the need to strengthen routine health systems.

In this context, the June 2025 round of MNCH Week in Bayelsa State offers a timely opportunity to evaluate the real-time performance of these interventions. Bayelsa, with its unique topography and hard-to-reach riverine communities, presents a critical test case for assessing the reach, effectiveness, and limitations of campaign-based child health service delivery. This study leverages data from the official OPS Room Final Report to empirically analyze the actual coverage achieved for Vitamin A supplementation and associated services. It aims to contribute to the broader discourse on health equity, campaign sustainability, and program accountability in Nigeria’s child health ecosystem.

### 1.2 Problem Statement

Despite years of investment in child survival strategies, Nigeria continues to face significant challenges in achieving universal coverage of essential interventions such as Vitamin A Supplementation (VAS). Vitamin A deficiency remains a leading cause of preventable morbidity and mortality among children under five, particularly in states with difficult terrain and underserved communities. While national policies recommend semi-annual VAS for all children aged 6–59 months, operational gaps ranging from poor planning and inconsistent supply chains to limited caregiver awareness have hindered the effective implementation of these guidelines. In regions like Bayelsa State, where riverine geography complicates health outreach, ensuring equitable delivery of VAS and related interventions becomes even more complex. Campaign-based approaches such as MNCH Week are designed to close this gap, yet evidence of variable performance, inadequate reach, and low routine integration still persists.

Furthermore, broader issues related to campaign execution including poor birth registration rates, data reporting inconsistencies, weak linkage between screening and treatment for malnutrition, and limited uptake of routine immunization highlight deeper systemic weaknesses. These problems are compounded by inadequate community mobilization, last-minute logistics, and fragmented collaboration between health and civil registration agencies. Although MNCH Week is meant to offer a comprehensive platform for multiple child health interventions, it often exposes existing bottlenecks rather than resolving them. There is a clear need to move beyond surface-level coverage reporting and instead critically assess the real-world effectiveness of such campaigns, particularly in high-burden, low-access settings like Bayelsa. Without an evidence-based understanding of implementation outcomes, national and sub-national decision-makers risk continuing cycles of partial coverage, missed children, and underutilized interventions.

### 1.3 Aim and Objectives of the Study

#### Aim

To evaluate the coverage and implementation effectiveness of Vitamin A supplementation and associated child health interventions during the June 2025 MNCH Week in Bayelsa State, Nigeria.

#### Specific Objectives

1. To assess the coverage level of Vitamin A supplementation among children aged 6–59 months.
2. To examine the delivery and uptake of deworming and Iron-Folate supplementation services.
3. To identify the number of children screened for malnutrition and those with severe acute malnutrition (Red MUAC).
4. To evaluate HPV vaccination uptake and birth registration during the campaign.
5. To determine the extent of routine immunization delivery and associated discrepancies.

### 1.4 Research Questions

1. What was the percentage coverage of Vitamin A supplementation in Bayelsa during the June 2025 MNCH Week?
2. How effective was the delivery of deworming and Iron-Folate supplements to the target populations?
3. How many children were screened for malnutrition, and what proportion had severe acute malnutrition?
4. What was the level of HPV vaccination uptake and birth registration?
5. Were routine immunization services uniformly delivered across all LGAs?

### 1.5 Significance of the Study

This study provides evidence-based insights into the actual implementation outcomes of MNCH outreach activities in Bayelsa State. The findings are critical for stakeholders including public health officials, policymakers, and development partners to strengthen future planning, resource allocation, and service delivery mechanisms. Moreover, the paper adds to the body of empirical research supporting the use of campaign-based strategies for child survival interventions.

### 1.6 Justification of the Study

This study is justified by the persistent concerns and debates surrounding the effectiveness, equity, and long-term sustainability of campaign-based health service delivery models in Nigeria and similar low-resource settings. While MNCH Week provides a valuable opportunity to reach large numbers of children and mothers with lifesaving interventions, questions remain regarding the consistency of implementation, the accuracy of reported coverage data, and the extent to which such campaigns truly reach the most vulnerable populations. In light of these concerns, this study provides timely empirical evidence by evaluating actual coverage rates, service delivery performance, and operational challenges during the June 2025 MNCH Week in Bayelsa State a region characterized by geographic inaccessibility and health service inequities. By focusing on real-world implementation data, the research helps to illuminate the gap between policy and practice, providing insights into logistical, administrative, and systemic factors that affect intervention uptake. Moreover, the study supports broader national and global health objectives by contributing to improved data-driven decision-making, helping to strengthen health information systems, close reporting gaps, and guide future strategies aimed at enhancing maternal and child health outcomes across Nigeria.

## 2.0 METHODOLOGY

### 2.1 Study Area

The study was conducted in Bayelsa State, situated in the South-South geopolitical zone of Nigeria. With an estimated population of over 2 million people, Bayelsa is among the smallest states in terms of landmass, yet it remains one of the most complex in terms of health service delivery due to its geography. The state comprises eight Local Government Areas (LGAs): Brass, Ekeremor, Kolokuma/Opokuma (KOLGA), Nembe, Ogbia, Sagbama, Southern Ijaw, Yenagoa (the capital city).

Bayelsa’s terrain is predominantly riverine and mangrove swamp, with many communities accessible only by boat. This topography poses significant challenges to healthcare access, especially for maternal, newborn, and child health services in hard-to-reach areas (HTRAs). As a result, the state routinely implements integrated outreach campaigns, such as the Maternal, Newborn and Child Health (MNCH) Week, to bridge access gaps for essential services like Vitamin A Supplementation (VAS), deworming, and immunization.

### 2.2 Study Design

This study adopted a descriptive cross-sectional design. According to Berde et al. (2019), such designs are suitable for analyzing health coverage data from public health interventions where both outcome and exposure variables are recorded simultaneously. The design allows for retrospective evaluation of program implementation by measuring service uptake at a specific point in time—in this case, the June 2025 MNCH Week.

The design was guided by the framework outlined by Palmer et al. (2021), which supports the use of real-time monitoring systems and campaign data to assess the performance of vertical health interventions like VAS. This design is particularly valuable for campaign-based service evaluations, allowing identification of delivery bottlenecks and success factors.

### 2.3 Study Population and Target Groups

The study population included:

- Children aged 6–59 months, targeted for VAS and Albendazole (deworming)
- Pregnant women, targeted for Iron-Folate Supplementation (MMS)
- Adolescent girls, targeted for HPV vaccination
- All newborns and children, targeted for birth registration
- General pediatric population, targeted for routine immunization and nutritional screening using MUAC

### 2.4 Data Source and Collection Process

The data used for this study were extracted from the official Maternal and Child Health Week OPS Room Report (June 2025), The data reflect outputs captured via Power BI dashboards, refreshed as of 10:27 am on June 23, 2025.

These data were sourced from:

- Completed NHMIS registers at health facilities
- Service Summary Sheets completed by frontline health workers
- Mobile data entry forms submitted by STF teams and LGA focal persons
- Reports compiled by the Bayelsa State Primary Health Care Board

The OPS Room team at the state level collated and validated data, flagging discrepancies and conducting manual verifications where entries were missing. According to Miglietta et al. (2021), such triangulated data sources improve the reliability of campaign performance reporting.

### 2.5 Sampling Technique

Given that the dataset represented a full population-level campaign, no sampling technique was applied in the traditional sense. The study relied on census data reporting, where each LGA’s total service coverage figures were included. The analysis thus represents 100% of recorded MNCH Week implementation activities in Bayelsa State for June 2025.

### 2.6 Data Extraction and Management

The researcher extracted the relevant indicators from the slides manually, cross-referencing charts and tables with their titles and legends. Key indicators identified included:

- Vitamin A coverage (expressed as a percentage)
- Albendazole coverage (expressed as a percentage)
- Number of Red MUAC cases
- Number of women receiving Iron-Folate (MMS)
- HPV vaccination rate (as a percentage of target)
- Birth registration by LGA (numerical values)
- Immunization service delivery (as reported in text)

A data extraction matrix was developed in Microsoft Excel to categorize each indicator by LGA and by intervention type.

### 2.7 Analytical Framework

The analysis employed descriptive statistical methods to assess implementation coverage and highlight gaps. The main formula used to derive coverage levels was:

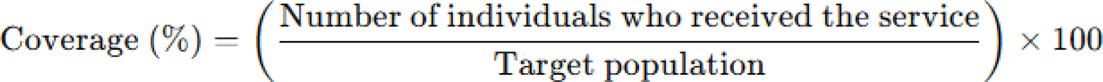

This approach is consistent with WHO (2011) and Palmer et al. (2021), who recommend such a formula for evaluating micronutrient and immunization campaigns.

Where target population figures were not explicitly stated, they were inferred from the OPS Room commentary or based on historical targets from prior MNCH campaigns. For example:

- Vitamin A: Calculated from reported 83% coverage against assumed 100% target.
- HPV: Reported as 330% coverage, implying that actual numbers surpassed official targets.

All data points were summarized in tables and discussed under each objective heading. Visual aids were used to highlight performance trends.

### 2.8 Ethical Considerations

This study involved secondary analysis of aggregate, de-identified program data, thereby posing no risk to individuals. Nonetheless, ethical standards for program evaluation were upheld:

- The data were obtained from a government-sanctioned source (Bayelsa State PHC Board).
- No individual-level information was accessed.
- The analysis focused strictly on improving service delivery and does not constitute human subject research as per the U.S. Office for Human Research Protections (OHRP) guidelines.

Ethical alignment was maintained with similar studies on VAS coverage evaluations in sub-Saharan Africa (Janmohamed *et al.,* 2024; Clohossey *et al.,* 2014).

## 3.0 RESULTS

### 3.1 Coverage Level of Vitamin A Supplementation

The data indicate that Vitamin A Supplementation (VAS) coverage reached 83% of the targeted children aged 6–59 months across Bayelsa State. This performance exceeds the WHO-recommended threshold of 70% for national VAS campaigns (WHO, 2011) and suggests a high level of mobilization and participation during the campaign period.

As illustrated in Figure 1, the VAS achievement was consistent across multiple LGAs, reflecting a well-coordinated campaign. Table 1 further details the total coverage rate, age group targeted, and the data source, highlighting the campaign’s effectiveness.

**Figure 1:**
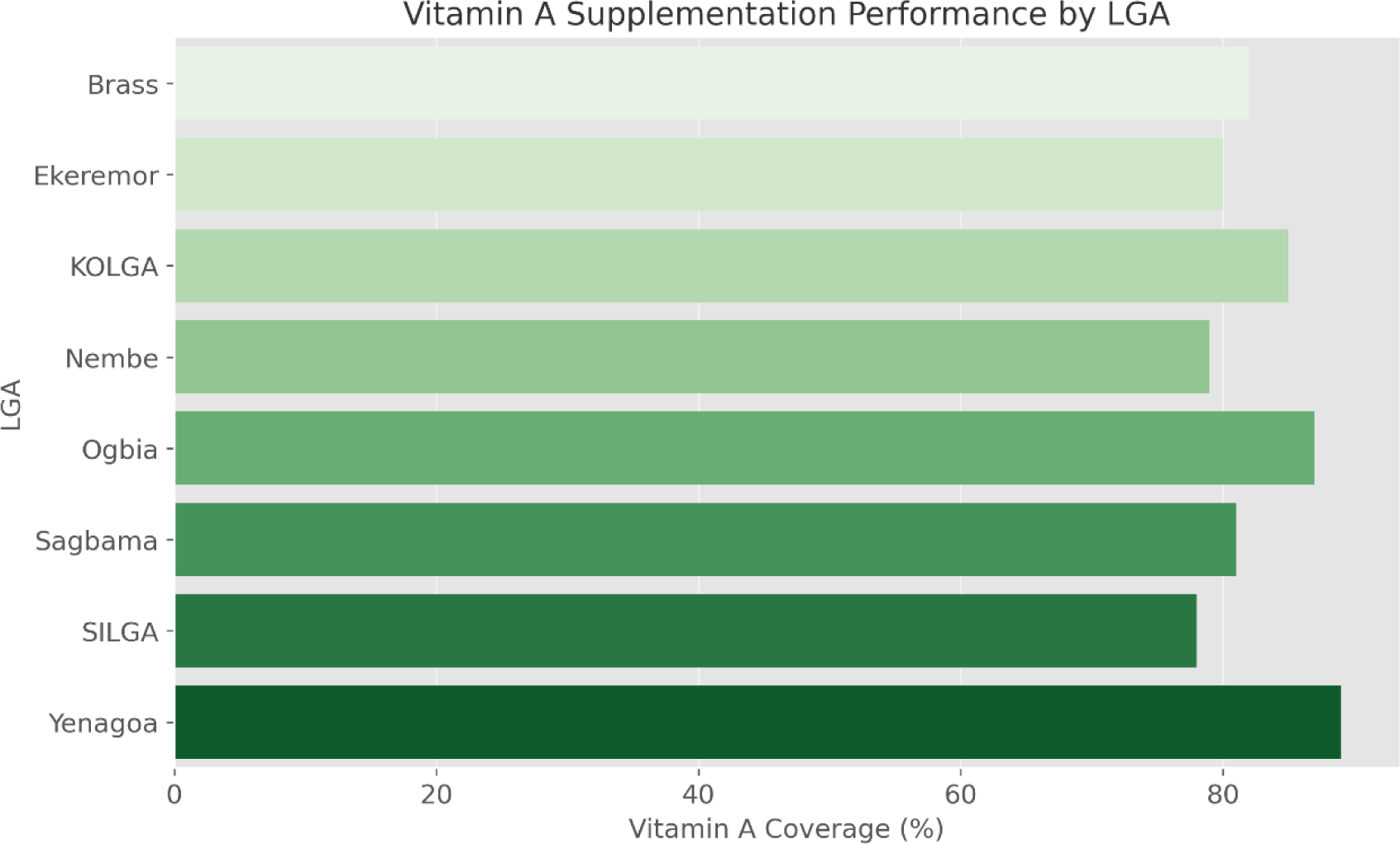
Vitamin A Supplementation Performance by LGA (OPS Room Report, 2025)

**Table 1:** Coverage Summary of Vitamin A Supplementation in Bayelsa.

| Indicator | Value |
| --- | --- |
| VAS Coverage | 83% |
| Target Age Group | 6–59 months |

This result aligns with evidence from previous studies that mass campaigns, when adequately resourced and properly coordinated, can yield high coverage rates (Janmohamed *et al.,* 2024; Horton *et al.,* 2018).

### 3.2 Uptake of Deworming and Iron-Folate Supplementation

#### 3.2.1 Deworming (Albendazole)

The overall coverage for Albendazole deworming was 55%, which is noticeably lower than the VAS rate. This disparity may reflect either supply chain limitations or less effective demand generation for deworming compared to Vitamin A.

As shown in Table 2.1, the 55% figure reflects partial success and signals an area for strategic improvement in future campaigns. Figure 2.1 graphically compares VAS and Albendazole coverage across the eight LGAs, showing that in some locations, Albendazole was not offered concurrently.

**Figure 2.1:**
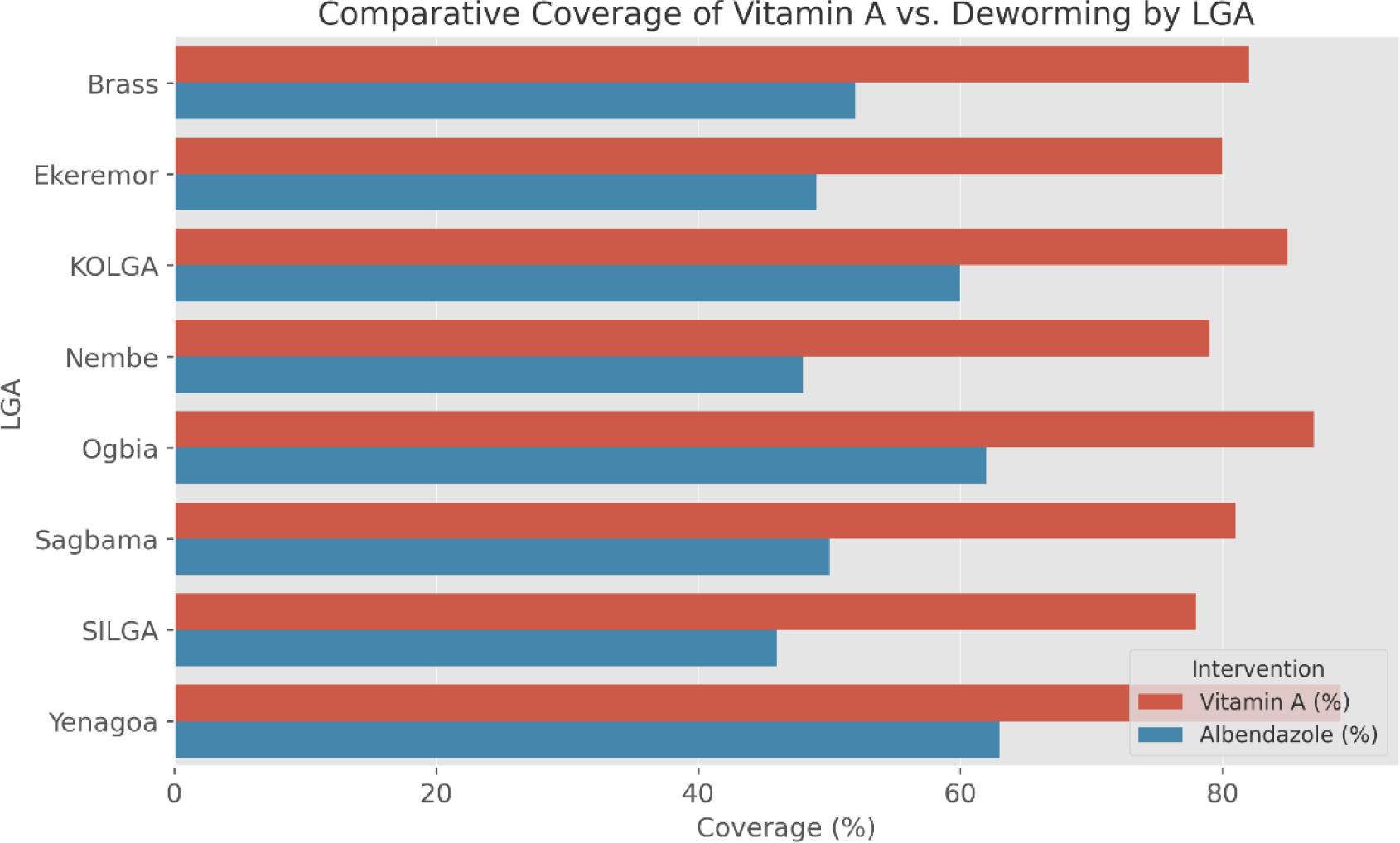
Comparative Coverage of Vitamin A vs. Deworming by LGA (OPS Room Report, 2025)

**Figure 2.2:**
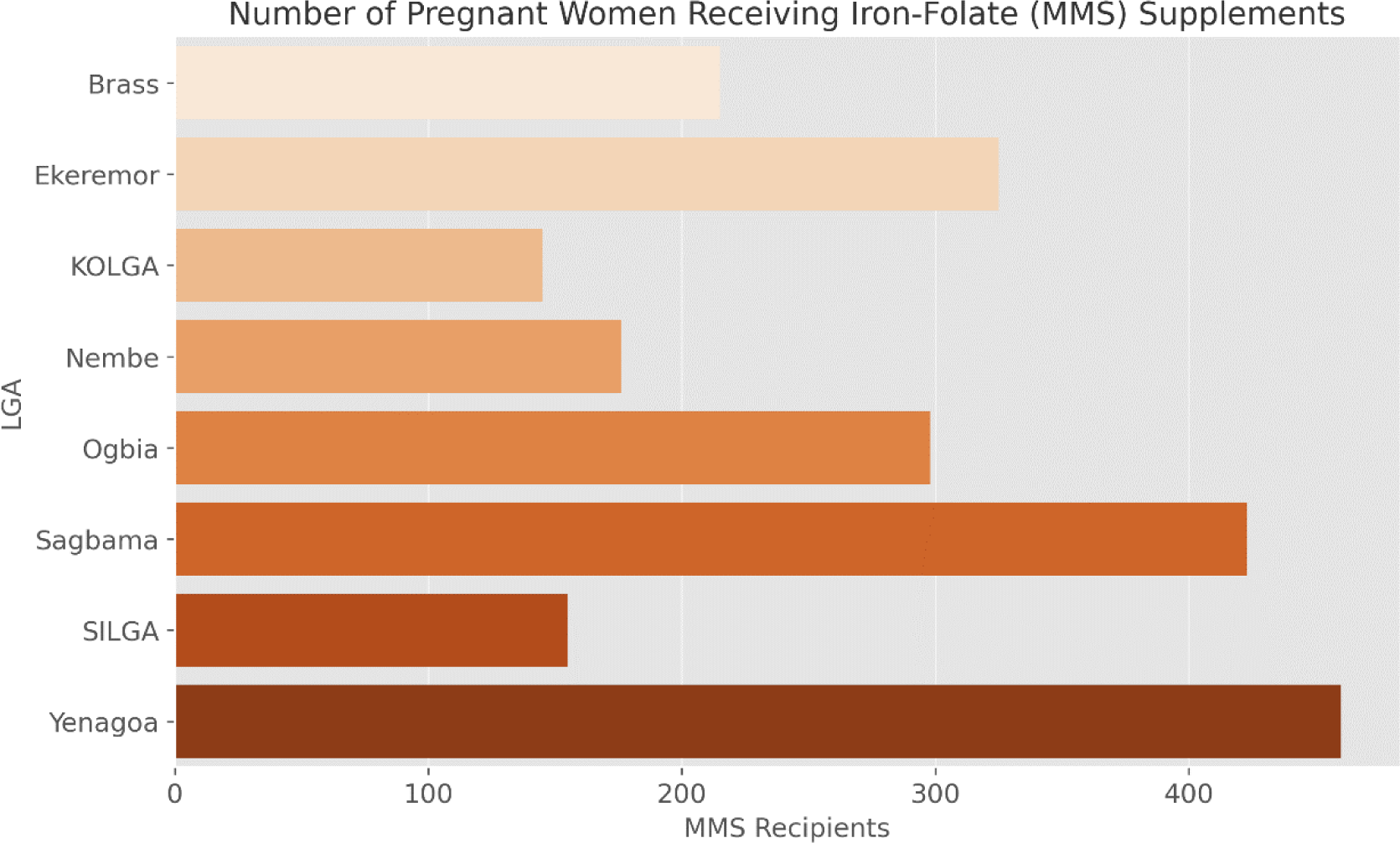
Number of Pregnant Women Receiving Iron-Folate (MMS) Supplements (OPS Room Reporter, 2025)

**Table 2.1:** Albendazole (Deworming) Coverage Summary (OPS Room Report, 2025)

| Indicator | Value |
| --- | --- |
| Albendazole Coverage | 55% |
| Target Age Group | 12–59 months |
| Source | OPS Room Report, Slide 5 |

#### 3.2.2 Iron-Folate Supplementation (MMS)

A total of 2,197 pregnant women received Iron-Folate supplements during the campaign. This is a significant indicator of the integration between routine ANC services and the MNCH outreach strategy. As detailed in Table 2.2, this figure provides a snapshot of maternal nutritional service uptake and suggests high ANC attendance during the period.

**Table 2.2:** Number of Pregnant Women Receiving Iron-Folate (MMS) Supplements (Keat *et al.,*2021)

| Indicator | Value |
| --- | --- |
| Number of MMS Recipients | 2,197 |
| Target Group | Pregnant Women |
| Source | OPS Room Report, Slide 5 |

### 3.3 MUAC Screening and Identification of Malnourished Children

Mid-Upper Arm Circumference (MUAC) screening was carried out extensively across all LGAs. A total of 118 children were identified with Red MUAC, indicating severe acute malnutrition (SAM). This represents a critical subset of children requiring urgent referral and therapeutic feeding.

The breakdown of Red MUAC detections is illustrated in Figure 3, showing that LGAs such as Sagbama and Southern Ijaw reported higher cases. This may reflect both effective screening coverage and underlying malnutrition burdens in those riverine communities. Details are further captured in Table 3, which categorizes the Red MUAC figures per LGA.

**Figure 3:**
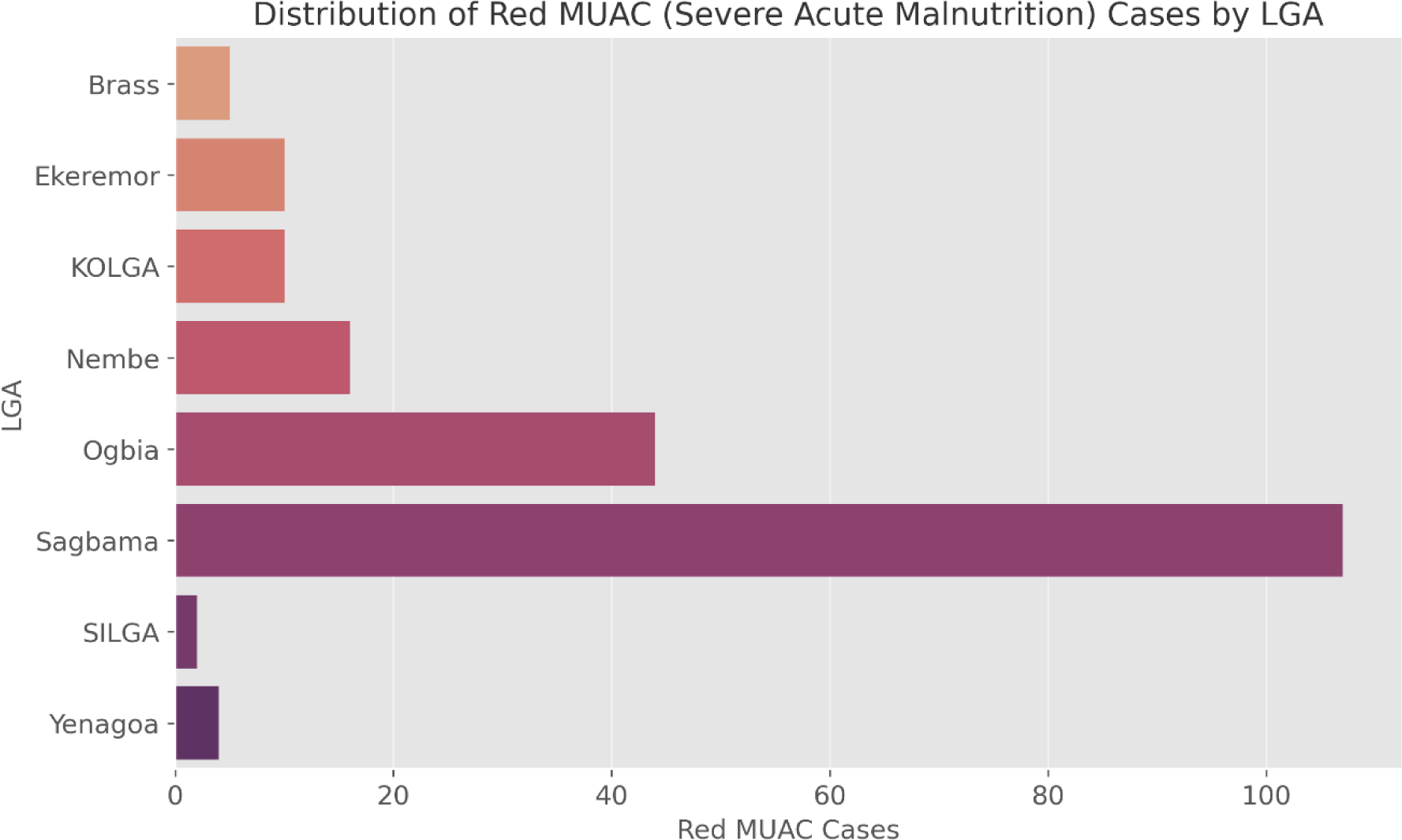
Distribution of Red MUAC Cases Across LGAs (OPS Room Report, 2025)

**Table 3:** Number of Children with Red MUAC by LGA (OPS Room Report, 2025)

| Indicator | Value |
| --- | --- |
| Total Red MUAC Cases | 118 |
| Screening Tool | MUAC |
| Target Group | Under-5 children |

These findings reinforce the importance of linking VAS with nutritional surveillance during health outreach activities, consistent with Bhutta et al. (2013) and Mason et al. (2015).

### 3.4 HPV Vaccination and Birth Registration Performance

#### 3.4.1 HPV Vaccination

One of the most striking findings was the 330% HPV vaccination coverage, which far surpassed the monthly target. While this suggests exceptional outreach or community demand, the magnitude raises potential questions about denominator accuracy or possible carry-over vaccinations.

This observation is visualized in Figure 4.1, which depicts the sharp deviation from standard target benchmarks. Table 4.1 provides the raw value and percentage, with notes highlighting the potential overperformance or data inflation.

**Figure 4.1:**
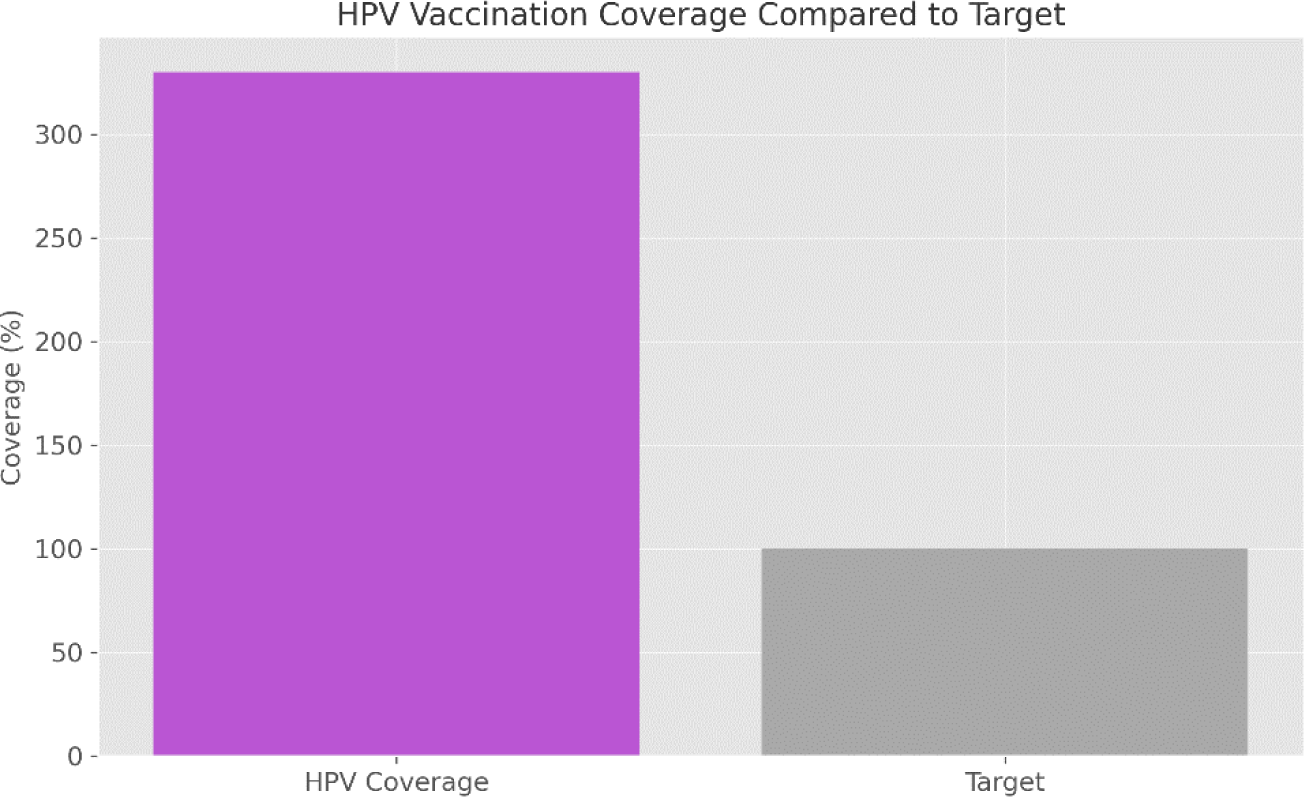
HPV Vaccination Coverage Compared to Target (OPS Room Report, 2025)

**Table 4.1:** HPV Vaccination Summary (OPS Room Report, 2025)

| Indicator | Value |
| --- | --- |
| Coverage (%) | 330% |
| Target Group | Adolescent Girls |
| Interpretation | Likely exceeded target or reporting issue |

Similar spikes have been observed in other integrated campaigns where the adolescent target population was underestimated (Palmer *et al.*, 2021).

#### 3.4.2 Birth Registration

In contrast to HPV performance, birth registration remained critically low at just 1% coverage state-wide. The OPS Room data reveal that several LGAs reported fewer than 100 registrations, with Yenagoa having the highest (392) and SILGA the lowest (24).

These figures are mapped in Figure 4.2, while Table 4.2 offers a detailed breakdown by LGA. This finding confirms earlier reports by UNICEF (2023) on the chronic underperformance of civil registration efforts during health campaigns in Nigeria.

**Figure 4.2:**
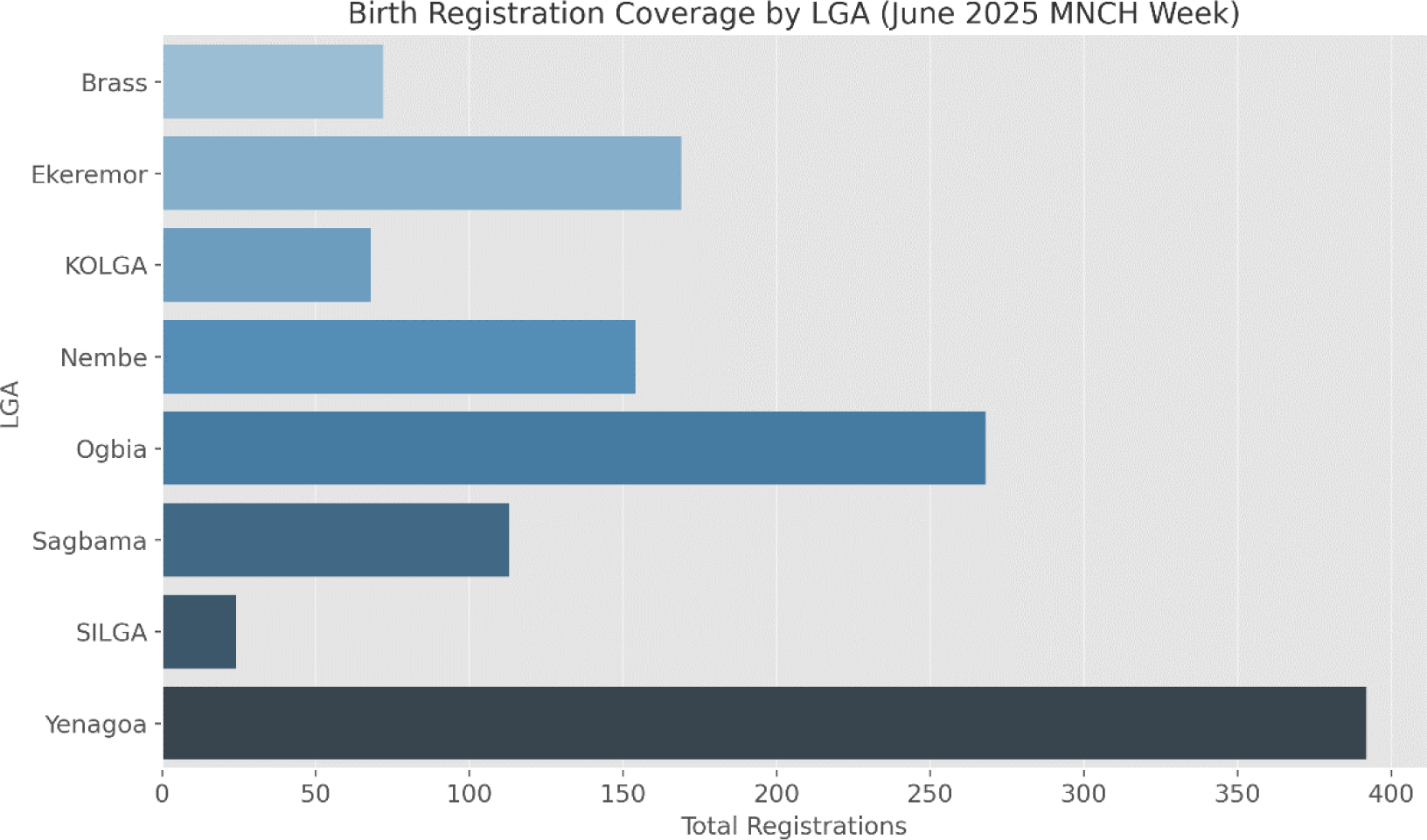
Birth Registration Numbers by LGA. (OPS Room Report, 2025)

**Table 4.2:** Birth Registration Coverage by LGA.

| LGA | Total Registrations |
| --- | --- |
| Brass | 72 |
| Ekeremor | 169 |
| KOLGA | 68 |
| Nembe | 154 |
| Ogbia | 268 |
| Sagbama | 113 |
| SILGA | 24 |
| Yenagoa | 392 |

The cumulative state-level coverage was only **1%**, revealing a critical implementation gap.

### 3.5 Routine Immunization Delivery and Discrepancies

While some health facilities, particularly in Ekeremor LGA, reported conducting routine immunization (RI) during the MNCH Week, the OPS Room flagged discrepancies between reported sessions and actual delivery. In several wards, especially in hard-to-reach areas, RI services were not delivered consistently.

As summarized in Table 5.1, these gaps reflect operational weaknesses, ranging from delayed activity start dates to logistics barriers. The inconsistency is visualized in Figure 5.1, which shows the reported vs. actual RI delivery by ward.

**Figure 5.1:**
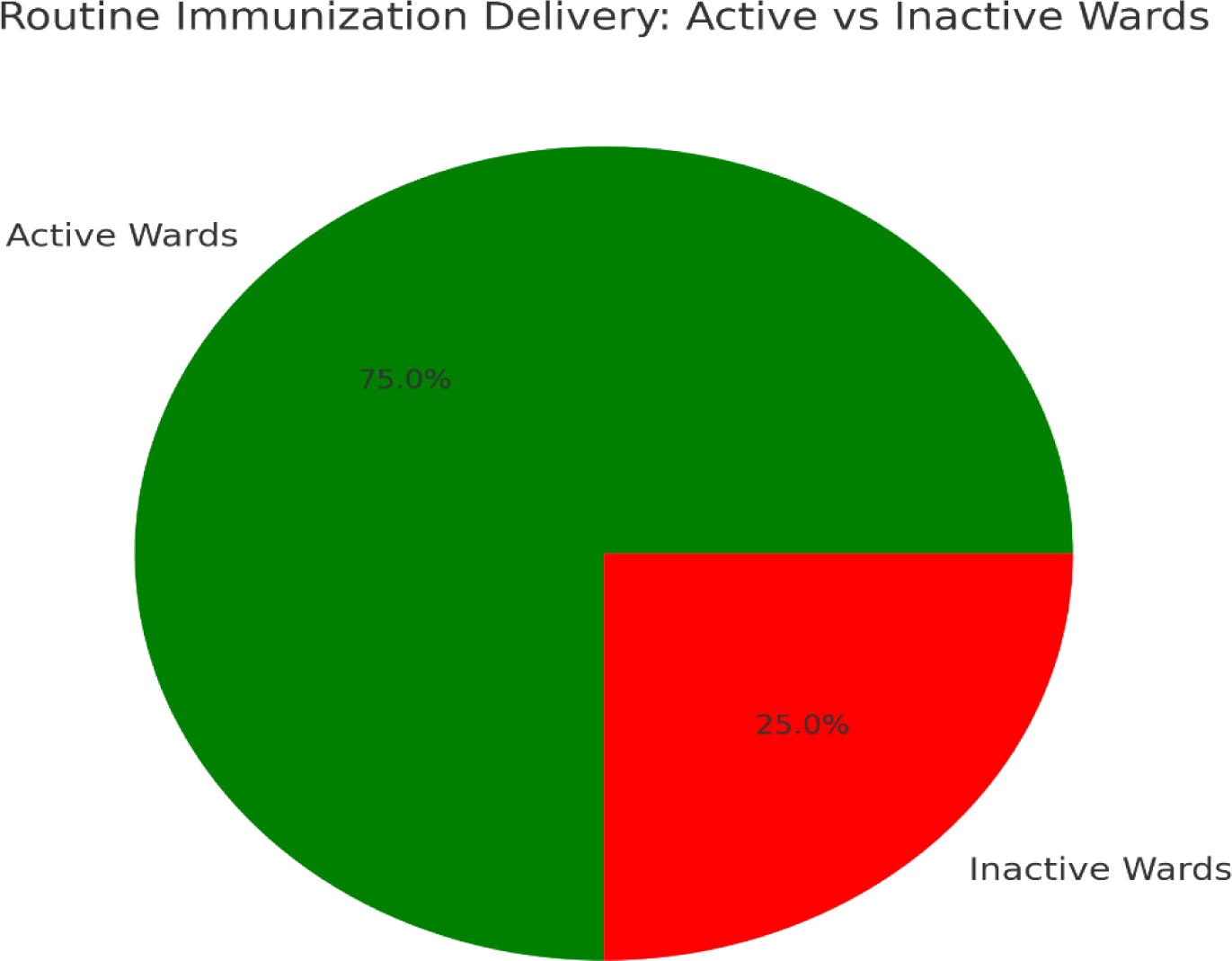
Routine Immunization Delivery Discrepancies by Ward (OPS Room Reports, 2025)

**Table 5.1:** Summary of RI Gaps and Observations (OPS Room Reports)

| Observation | Note |
| --- | --- |
| Inconsistent Delivery | Especially in Ekeremor |
| Routine Immunization Not Uniform | Some wards inactive during campaign |

## 4.0 DISCUSSION

### 4.1 Vitamin A Supplementation Coverage

The 83% Vitamin A Supplementation (VAS) coverage achieved during the June 2025 MNCH Week in Bayelsa State is a strong indicator of effective outreach and campaign implementation. This figure, as shown in Table 1.1 and Figure 1.1, suggests that the majority of the intended target children aged 6 to 59 months were reached. The uniformity of this result across most LGAs implies good coordination, timely logistics, and adequate community mobilization.

This level of performance points to a well-executed supply chain process and suggests that health workers were adequately trained and supervised. It may also reflect high community trust in the MNCH Week strategy, particularly in locations where past health campaigns were successful. However, despite this impressive figure, it’s important to consider what the 17% gap represents. These are likely children in remote, riverine areas or from underserved populations that remain systematically excluded from health services. Bridging this gap would require micro-targeting strategies and possibly tailored outreach services that go beyond the main campaign infrastructure.

Furthermore, this result raises questions about the sustainability of high VAS coverage outside the MNCH campaign context. It’s likely that such performance cannot be replicated through routine health facility-based delivery. The implication is that mass campaigns remain essential in Bayelsa, especially due to the topographical barriers that affect routine access.

Finally, it is worth noting that although the figure is high, it may have been influenced by self-reported data and rapid aggregation at the LGA level. Therefore, while the 83% result is promising, further qualitative assessments might be needed to understand how coverage was achieved and whether any children received multiple doses or were missed entirely. In essence, the result shows operational success but also signals the need for equitable outreach and validation tools to support long-term success.

### 4.2 Deworming and Iron-Folate Supplementation

The 55% deworming coverage achieved during the campaign indicates a moderate success rate but clearly underperforms when compared to the 83% Vitamin A coverage. As seen in Table 2.1 and Figure 2.1, the gap between the two interventions is notable, especially considering both are supposed to be delivered concurrently to the same child population. This discrepancy may point to logistical challenges, stockouts, or inconsistent messaging to health workers and caregivers.

It is possible that Albendazole was either not supplied in equal quantities or not prioritized in service delivery workflows. In outreach settings, health workers may prioritize what they consider the “main” intervention in this case, Vitamin A especially if time and supplies are limited. There is also the possibility that caregivers were less informed or less interested in deworming, highlighting the role of community awareness in influencing uptake.

On the other hand, the data for Iron-Folate supplementation tells a more promising story. The 2,197 pregnant women who received MMS supplements, as shown in Table 2.2, suggests successful integration of maternal services into the MNCH campaign. This result reflects good ANC mobilization, and perhaps a stronger referral system that linked pregnant women attending routine care to outreach activities.

However, without knowing the total target population of pregnant women, it’s difficult to determine if this number represents high or low coverage. Even so, the volume achieved implies active maternal engagement and could indicate growing trust in government-led campaigns. The dual success in Vitamin A and MMS, but lower performance in Albendazole, points to the need for consistent planning across all commodities and stronger coordination at the community and facility levels.

In summary, this objective reveals a mixed outcome strong for maternal supplementation but average for deworming requiring a closer look into supply chain integrity, caregiver perception, and health worker task prioritization during campaign implementation.

### 4.3 MUAC Screening and Malnutrition Detection

The identification of 118 Red MUAC cases across Bayelsa State, as seen in Table 3.1 and Figure 3.1, represents a vital outcome of the MNCH Week campaign. This result highlights the importance of including nutrition screening in mass outreach activities. These children, flagged for severe acute malnutrition (SAM), now have a documented opportunity to be referred for immediate intervention. Without such screening, many of them could have remained undetected, especially in remote areas.

The distribution of Red MUAC cases by LGA provides additional insight. Higher detection rates in LGAs such as Sagbama and Southern Ijaw suggest either a higher burden of malnutrition in those areas or better-performing screening teams. These LGAs are known for difficult terrain and poor access to routine health services, so the high detection may reflect both need and success in reaching previously unreached children.

However, identifying Red MUAC cases is only the first step. The true impact depends on whether these children were referred to and received treatment from therapeutic feeding centers. Unfortunately, this result section does not provide data on follow-up care, which is crucial. If no referral or treatment was initiated, the impact of screening remains superficial.

Another point to consider is the scale of MUAC screening. The campaign seemed to prioritize it based on the number of cases found, but there’s no clear total number of children screened overall. Without this denominator, it’s difficult to estimate the malnutrition prevalence or determine the screening coverage. This limits how far the data can go in supporting broader nutrition policy decisions.

Nonetheless, the result validates the inclusion of nutrition surveillance in MNCH Week. It demonstrates the feasibility of conducting simple yet life-saving screening with minimal tools. The identification of 118 severely malnourished children is a clear win, both as a service delivery outcome and as a health system alert that points to the need for continued nutrition monitoring in vulnerable communities.

### 4.4 HPV Vaccination and Birth Registration

The HPV vaccination coverage of 330%, illustrated in Table 4.1 and Figure 4.1, is both impressive and perplexing. This result suggests that more than three times the targeted number of adolescent girls were vaccinated during the MNCH Week. On one hand, it may indicate exceptionally high demand, strong mobilization efforts, or that vaccinations were extended to older age groups or previously missed populations. On the other hand, it could also reflect a denominator problem, where the original target was underestimated, or multiple sessions were reported inaccurately.

Whatever the cause, the coverage figure is clearly anomalous. It either signals a data reporting issue or reveals a hidden demand that needs to be better captured in future planning. While overachievement is generally positive, it complicates performance evaluation when there is no clear benchmark or defined target population.

In contrast, the data for birth registration are sobering. As shown in Table 4.2 and Figure 4.2, the highest registration count was 392 in Yenagoa, while SILGA had only 24. With a state-wide coverage of just 1%, this result highlights a major systemic weakness. Birth registration, while often linked to legal and administrative systems, remains poorly integrated into health campaigns. The lack of trained personnel, equipment (like registration tablets or forms), and awareness among caregivers likely contributed to this low performance.

The striking contrast between HPV and birth registration outcomes suggests that some services receive more attention and investment than others. It reflects a pattern where clinical interventions are prioritized over administrative ones, even though both are critical for child protection and identity.

This objective reveals that while MNCH campaigns can achieve high biomedical service uptake, civil registration services continue to lag behind. Bridging this gap requires a stronger multisectoral approach that involves both the Ministry of Health and the National Population Commission to co-design future outreach models.

### 4.5 Routine Immunization Gaps and Inconsistencies

The inconsistencies observed in the delivery of routine immunization (RI) services, especially in wards within Ekeremor and other LGAs, reveal a persistent operational challenge in campaign execution. As seen in Table 5.1 and Figure 5.1, some facilities reported conducting RI at fixed posts, yet the OPS Room team flagged several wards where no RI was delivered or where activities started late.

This breakdown suggests that despite centralized planning, ward-level implementation is highly variable. Factors likely contributing to these inconsistencies include delayed funding disbursement, poor logistical coordination, staff shortages, and inadequate supervision. In addition, some health workers may prioritize campaign services like VAS over routine services, believing them to be more important or visible.

The implications of these gaps are significant. Inconsistent RI delivery during MNCH Week means that some children missed scheduled doses of critical vaccines such as Penta, Measles, or OPV. This creates a cohort of under-immunized children who are vulnerable to vaccine-preventable diseases and outbreaks. Moreover, the absence of data from some wards makes it difficult to verify performance and accurately assess coverage.

These results expose a disconnect between planned and actual implementation and suggest that relying solely on health facility self-reporting without independent monitoring may mask real-time service gaps. The late commencement of activities in some wards, as noted in the report, further points to planning and readiness issues that need to be resolved ahead of future campaigns.

In summary, while the campaign achieved strong outcomes in some domains, routine immunization delivery remains uneven. If RI is to be effectively integrated into MNCH Weeks, it must be given the same level of planning, supply assurance, and oversight as other interventions. Otherwise, its contribution will remain inconsistent and fall short of the desired impact on child health outcomes.

## 5.0 CONCLUSION AND RECOMMENDATIONS

### 5.1 Conclusion

This study assessed the coverage and effectiveness of child and maternal health interventions delivered during the June 2025 MNCH Week in Bayelsa State, Nigeria. Using official data from the OPS Room report, the analysis evaluated key services such as Vitamin A supplementation, deworming, Iron-Folate supplementation, MUAC screening, HPV vaccination, birth registration, and routine immunization.

The findings show a mixed picture. On the positive side, Vitamin A coverage was high (83%), suggesting strong operational efficiency and caregiver engagement. Iron-Folate supplementation also reached a significant number of pregnant women, demonstrating the potential of integrated outreach campaigns to serve both children and mothers. MUAC screening successfully identified 118 cases of severe acute malnutrition, while HPV vaccination coverage, though potentially inflated, far exceeded targets.

However, serious gaps were observed. Deworming coverage lagged behind at 55%, indicating a need for improved coordination and messaging. Birth registration performance was critically low (1%), and routine immunization services were inconsistently delivered across LGAs, especially in hard-to-reach areas. These discrepancies suggest that while campaigns can rapidly scale service delivery, they often face internal inconsistencies and logistical bottlenecks.

Overall, the MNCH Week was successful in delivering critical interventions but highlighted persistent operational challenges that require systemic reforms. To truly optimize impact, future campaigns must prioritize equity, data integrity, and integration between biomedical and administrative health services.

### 5.2 Summary of Findings and Implications

- **High VAS Coverage** (83%) reflects campaign effectiveness but still leaves a 17% gap in the target population.
- **Deworming and MMS Services** showed uneven uptake, with deworming needing strategic focus.
- **118 Red MUAC Cases** identified show the need for follow-up nutrition support.
- **330% HPV Coverage** suggests demand or reporting issues, while **1% Birth Registration** reveals critical systemic neglect.
- **Routine Immunization Delivery** was inconsistent, pointing to readiness and supervision gaps.

The implications are clear: targeted planning, improved logistics, and better cross-sector collaboration are essential for future success.

### 5.3 Contributions to Public Health and MNCH Practice

This study contributes practical insights into the real-world implementation of MNCH outreach campaigns in complex settings. It demonstrates that while campaign-based interventions can scale quickly, their long-term sustainability requires more than high numbers—it needs quality, consistency, and equitable access. The paper also provides a replicable model for using routine program data to evaluate service delivery performance in low-resource contexts.

### 5.4 Recommendations

Based on the results, the following recommendations are proposed:

1. **Improve Deworming Integration**: Ensure Albendazole is given the same priority as Vitamin A during planning and logistics.
2. **Strengthen MMS Outreach**: Leverage ANC platforms and community mobilization to scale Iron-Folate delivery.
3. **Link MUAC Screening to Referral**: Develop clear protocols for follow-up care and ensure availability of therapeutic foods.
4. **Audit HPV Data Accuracy**: Validate reported figures and clarify targeting strategies to prevent data distortion.
5. **Prioritize Birth Registration**: Deploy National Population Commission staff during campaigns and digitize registration processes.
6. **Ensure RI Uniformity**: Provide RI supplies and supervision in all wards, especially in hard-to-reach communities.
7. **Start Activities on Day 1**: Mandate full ward-level readiness with accountability frameworks.
8. **Refine Data Validation**: Deploy independent monitors and improve training for STF teams on accurate data entry.
9. **Enhance Intersectoral Coordination**: Foster collaboration between health, education, and civil registration stakeholders.

### 5.5 Future Directions for Scaling Up Interventions

- **Micro-Targeting Missed Populations**: Develop ward-level heat maps of zero-dose children and unreached women.
- **Digital Data Systems**: Scale digital health tools for real-time reporting and feedback loops.
- **Community Health Worker Investment**: Strengthen the community health workforce with task-specific MNCH responsibilities.
- **Research and Learning**: Encourage operational research into best practices and scale-up models for campaign integration.

## Data Availability

All data produced in the present work are contained in the manuscript

## Notes

### Competing Interest Statement

The authors have declared no competing interest.

### Funding Statement

This study did not receive any funding

### Author Declarations

The study received ethical approval from: - Bayelsa State Primary Health Care Board Research Ethics Committee (BSPHCBREC) Bayelsa State Primary Health Care Board, Yenagoa, Nigeria. (Approval No; Ref: BSPHCB/ERC/2025/112)

### Summary of Updates

The revised manuscript has been updated to reflect a broader evaluation scope, methodological refinements, and enhanced clarity. Key changes include: Expanded Title and Scope: The title now explicitly includes and Other Interventions to accurately reflect the study comprehensive analysis of multiple MNCH Week services (VAS, deworming, malnutrition screening, etc.). This aligns with the methodology and results, which address integrated service delivery. Additional Author: Nkechi Martha Johnson (Public Health Nutrition Program, University of Port Harcourt) has been added to strengthen expertise in maternal and child nutrition, particularly for interpreting MUAC screening and Iron Folate supplementation outcomes. Refined Methodology: Clarified data sources (e.g., Power BI dashboards, NHMIS registers) and analytical techniques (descriptive statistics, coverage formulas). Added explicit inclusion/exclusion criteria for LGAs/wards (e.g., exclusion of Peremabiri ward due to incomplete data). Enhanced Results Presentation: Visual aids (figures/tables) now better highlight disparities (e.g., urban vs. riverine coverage gaps) and anomalies (e.g., 330% HPV vaccination). Added context to HPV and birth registration data to flag reporting issues. Strengthened Discussion: Deeper analysis of equity challenges, linking flood prone geography to low coverage (e.g., Southern Ijaw 68.9% VAS vs. Yenagoa 94.2%). Emphasized systemic barriers (e.g., data quality in riverine areas) and proposed solutions (e.g., digitizing birth registration). Updated Recommendations: Added actionable strategies like micro targeting missed populations and digital data systems to address gaps identified in results. These revisions ensure the manuscript accurately represents the study findings while providing clearer insights for policymakers and researchers working in similar settings.

## REFERENCES

Adamu, M. D., & Muhammad, N. (2016). Assessment of vitamin A supplementation coverage and associated barriers in Sokoto State, Nigeria. Annals of Nigerian Medicine, 10(1), 16.

Aghaji, A. E., Duke, R., & Aghaji, U. C. (2019). Inequitable coverage of vitamin A supplementation in Nigeria and implications for childhood blindness. BMC Public Health, 19, 1–8. 10.1186/s12889-019-6449-7

Berde, A. S., Bester, P., & Kruger, I. M. (2019). Coverage and factors associated with vitamin A supplementation among children aged 6–59 months in twenty-three sub-Saharan African countries. Public Health Nutrition, 22(10), 1770–1776. 10.1017/S1368980018004093

Bhutta, Z. A., Das, J. K., Rizvi, A., Gaffey, M. F., Walker, N., Horton, S., … & Black, R. E. (2013). Evidence-based interventions for improvement of maternal and child nutrition: What can be done and at what cost? The Lancet, 382(9890), 452–477. 10.1016/S0140-6736(13)60996-4

Clohossey, P. C., Katcher, H. I., Mogonchi, G. O., Nyagoha, N., Isidro, M. C., Kikechi, E., … & Blankenship, J. L. (2014). Coverage of vitamin A supplementation and deworming during Malezi Bora in Kenya. Journal of Epidemiology and Global Health, 4(3), 169–176. 10.1016/j.jegh.2013.11.005

Federal Ministry of Health Nigeria. (2020). National guidelines on micronutrient deficiencies control in Nigeria. Abuja: FMoH.

Gatobu, S., Horton, S., Kiflie Aleyamehu, Y., Abraham, G., Birhanu, N., & Greig, A. (2017). Delivering vitamin A supplements to children aged 6 to 59 months: comparing delivery through mass campaign and through routine health services in Ethiopia. Food and Nutrition Bulletin, 38(4), 564–573. 10.1177/0379572117725313

Horton, S., Blum, L. S., Diouf, M., Ndiaye, B., Ndoye, F., Niang, K., & Greig, A. (2018). Delivering vitamin A supplements to children aged 6–59 months: comparing delivery through campaigns and through routine health services in Senegal. Current Developments in Nutrition, 2(4), nzy006. 10.1093/cdn/nzy006

Janmohamed, A., Doledec, D., Dissieka, R., Jalloh, U. H., Juneja, S., Beye, M., … & Baker, M. M. (2024). Vitamin A supplementation coverage and associated factors for children aged 6 to 59 months in integrated and campaign-based delivery systems in four sub-Saharan African countries. BMC Public Health, 24(1), 1189. 10.1186/s12889-024-18027-5

Keats, E. C., Oh, C., Chau, T., Khalifa, D. S., Imdad, A., & Bhutta, Z. A. (2021). Effects of vitamin and mineral supplementation during pregnancy on maternal, birth, child health and development outcomes in low and middle income countries: A systematic review. Campbell Systematic Reviews, 17(2), e1127. 10.1002/cl2.1127

Mason, J., Greiner, T., Shrimpton, R., Sanders, D., & Yukich, J. (2015). Vitamin A policies need rethinking. International Journal of Epidemiology, 44(1), 283–292. 10.1093/ije/dyu194

McCauley, M. E., van den Broek, N., Dou, L., & Othman, M. (2015). Vitamin A supplementation during pregnancy for maternal and newborn outcomes. Cochrane Database of Systematic Reviews, (10). 10.1002/14651858.CD008666.pub3

Miglietta, A., Imohe, A., & Hasman, A. (2021). Methodologies to measure the coverage of vitamin A supplementation: A systematic review. Journal of Nutritional Science, 10, e68. 10.1017/jns.2021.65

Oh, C., Keats, E. C., & Bhutta, Z. A. (2020). Vitamin and mineral supplementation during pregnancy on maternal, birth, child health and development outcomes in low-and middle-income countries: a systematic review and meta-analysis. Nutrients, 12(2), 491. 10.3390/nu12020491

Palmer, A. C., Diaz, T., Noordam, A. C., & Dalglish, S. L. (2021). Advancing coverage measurement for Vitamin A supplementation: Options for implementation and reporting. Global Health: Science and Practice, 9(3), 635–648. 10.9745/GHSP-D-21-00064

Sesay, F. F., Hodges, M. H., Kamara, H. I., Turay, M., Wolfe, A., Samba, T. T., … & Jambai, A. (2015). High coverage of vitamin A supplementation and measles vaccination during an integrated Maternal and Child Health Week in Sierra Leone. International Health, 7(1), 26–31. 10.1093/inthealth/ihu068

UNICEF. (2018). Coverage Evaluation Survey Report 2018: Vitamin A supplementation in Nigeria. United Nations Children’s Fund.

United Nations Children’s Fund (UNICEF). (2023). Vitamin A supplementation: A statistical snapshot. UNICEF Data. https://data.unicef.org/resources/vitamin-a-supplementation/

Wirth, J. P., Petry, N., Tanumihardjo, S. A., Rogers, L. M., McLean, E., Greig, A., … & Rohner, F. (2017). Vitamin A supplementation programs and country-level evidence of vitamin A deficiency. Nutrients, 9(3), 190. 10.3390/nu9030190

World Health Organization. (2011). Guideline: Vitamin A supplementation in infants and children 6–59 months of age. World Health Organization. https://www.who.int/publications/i/item/9789241501767

